# Tracking changes in reporting of epidemiological data during the COVID-19 pandemic in Southeast Asia: an observational study during the first wave

**DOI:** 10.1101/2020.10.23.20217570

**Authors:** Arianna Maever L. Amit, Veincent Christian F. Pepito, Bernardo Gutierrez, Thomas Rawson

**Author notes:** Corresponding author: Arianna Maever L. Amit MAS, College of Medicine, University of the Philippines Manila, Manila, Philippines.

## Abstract

**Background:** When a new pathogen emerges, consistent case reporting is critical for public health surveillance. Tracking cases geographically and over time is key for understanding the spread of an infectious disease and how to effectively design interventions to contain and mitigate an epidemic. In this paper we describe the reporting systems on COVID-19 in Southeast Asia during the first wave in 2020, and highlight the impact of specific reporting methods.

**Methods:** We reviewed key epidemiological variables from various sources including a regionally comprehensive dataset, national trackers, dashboards, and case bulletins for 11 countries during the first wave of the epidemic in Southeast Asia. We recorded timelines of shifts in epidemiological reporting systems. We further described the differences in how epidemiological data are reported across countries and timepoints, and the accessibility of epidemiological data.

**Findings:** Our findings suggest that countries in Southeast Asia generally reported precise and detailed epidemiological data during the first wave of the COVID-19 pandemic. However, changes in reporting were frequent and varied across data and countries. Changes in reporting rarely occurred for demographic data such as age and sex, while reporting shifts for geographic and temporal data were frequent. We also found that most countries provided COVID-19 individual-level data daily using HTML and PDF, necessitating scraping and extraction before data could be used in analyses.

**Interpretation:** Countries have different reporting systems and different capacities for maintaining consistent reporting of epidemiological data. As the pandemic progresses, governments may also change their priorities in data sharing. Our study thus highlights the importance of more nuanced analyses of epidemiological data of COVID-19 within and across countries because of the frequent shifts in reporting. Further, most countries provide data on a daily basis but not always in a readily usable format. As governments continue to respond to the impacts of COVID-19 on health and the economy, data sharing also needs to be prioritised given its foundational role in policymaking, and the implementation and evaluation of interventions.

**Funding:** The work was supported through an Engineering and Physical Sciences Research Council (EPSRC) (https://epsrc.ukri.org/) Systems Biology studentship award (EP/G03706X/1) to TR. This project was also supported in part by the Oxford Martin School.

The funders had no role in study design, data collection and analysis, decision to publish, or preparation of the manuscript.

## Research in context

Ongoing research into the epidemiology of SARS-CoV-2 depends entirely on access to regularly updated, factor-rich data. The benefits and importance of data-sharing practices have been well-documented during previous outbreaks, however the scale of the global pandemic currently faced presents its own unique challenges. The majority of countries are routinely reporting the number of confirmed cases and deaths attributed to COVID-19, with the country-wide cumulative totals readily accessible from databases such as the one curated by Johns Hopkins University.^9^ However, the breadth of further information reported by each country is less understood. Access to demographic and geographic information of cases in particular is critically important in the context of informing response policy, as these provide greater insights into how subgroups of the population in different areas are affected by the disease. Understanding how and when these data is provided is critical to ensuring that modelling efforts and government response are well-informed.

### Evidence before this study

The current research into the quality of data reporting is severely limited, with studies focussing primarily around specific locations, time periods, or population sub-groups. One research group has examined the data availability for 507 patients reported in January, finding that the majority of information was provided by social media and news outlets. Other than this example, there is no other work that we are aware of that investigates the issues surrounding data availability. Our work is the first to explore the scale of data reporting across the broader pandemic timeline.

### Added value of this study

Our research provides wider insight into the data pipeline from government to researchers, and how it has adapted over time. This timeline provides greater context to the specific findings of subsequent data-driven research, highlighting areas and time periods where particular data feeds are likely to be particularly biased or data-sparse. We are also able to recommend, based upon our findings, prioritising the use of the early-case histories of specific countries for the calculation of demographic-specific disease parameters. By highlighting particular regions where specific data are available, such as travel history, hospitalisation times and symptom-tracking, we are also able to identify ideal further topics of research in the ongoing attempts to fight the spread of COVID-19.

### Implications of all the available evidence

We reviewed the data of the Open COVID-19 Data Working Group’s centralised repository and other relevant data sources to compare the reporting systems of 11 Southeast Asian countries. We found that governments frequently changed the type of data and level of detail reported as the pandemic progressed. Our study thus highlights the importance of more nuanced analyses of epidemiological COVID-19 data within and across countries because of the frequent shifts in reporting. Further, most countries provide data on a daily basis but not always in a readily usable format. As governments continue to respond to the impacts of COVID-19 on health and the economy, data sharing also needs to be prioritised given its foundational role in policymaking, and the implementation and evaluation of interventions.

## Introduction

In December 2019, an outbreak of severe acute respiratory syndrome coronavirus 2 (SARS-CoV-2) was reported in Wuhan, China and was determined to cause the novel coronavirus disease 2019 (COVID-19). The World Health Organization (WHO) declared the outbreak to be a Public Health Emergency of International Concern on 30 January 2020, and subsequently a pandemic on 11 March 2020.

To effectively respond to public health emergencies, there is a need for timely and accurate reporting of statistics and data sharing as highlighted in the recent Ebola and Zika epidemics.^1-4^ To this end, the Principles for Data Sharing in Public Health Emergencies consisting of timeliness, ethics, equitability, accessibility, transparency, fairness, and quality have been developed and introduced.^3,5,6^ As of writing, only one study on data sharing during disease outbreaks among Southeast Asian countries has been carried out.^7^ The study evaluated data quality and timeliness of outbreak reporting in Cambodia, Lao PDR, Myanmar, and Vietnam for dengue, food poisoning and diarrhea, severe diarrhea, diphtheria, measles, H5N1 influenza, H1N1 influenza, rabies, and pertussis. Further, it highlighted the broad differences observed in the data quality and timeliness between participating countries, concluding that any international data-curating attempts must be versatile enough to accomodate this.

In the ongoing COVID-19 crisis, government organisations, public health agencies, and research groups are responding to the call for rapid data sharing by providing data and curating detailed real-time databases that are readily and publicly accessible.^8–11^ Data from various groups have informed more than 100,000 papers on COVID-19.^12^ Despite progress in reporting and sharing data, several challenges remain. First, there are ethical and privacy considerations that need to be balanced carefully against the potential impact of open data sharing. Second, there is a clear lack of capacity and often appropriate computational infrastructure that may make data sharing in real time unfeasible and burdensome.^3,4^ Such challenges may result in changes in the quality and detail of data reporting between and within countries over time as their respective health systems become increasingly overwhelmed.^11^ These shifts in reporting provide a challenge for accurately comparing epidemiological situations between countries. In China, for example, it has been shown that changes in reporting have impacted modelling results of the transmission parameters of COVID-19.^13^ Further, as the pandemic progresses and epidemiological information becomes increasingly less available, analyses of detailed case counts that cover the entire duration of the epidemic may not be feasible.^10^

To our knowledge, this is the first study to describe the ways in which various countries in a geographic region report COVID-19 data, and how the detail of data reporting changed over time. We reviewed detailed epidemiological data from Southeast Asian countries and tracked how countries’ reporting of COVID-19 data has shifted. We further evaluated differences in reporting between countries and described the accessibility of epidemiological data during the first wave in 2020. This study is descriptive and does not seek to evaluate health systems reporting. However, by providing these types of information, researchers may be able to conduct better and more nuanced analyses of epidemiological data of COVID-19. By showing changes in reporting, we hope to provide insights on how data should be viewed and analysed.

## Methods

### Study design

We conducted an observational study to describe and track changes in reporting of epidemiological data during the COVID-19 pandemic in 11 countries in Southeast Asia, namely Brunei, Cambodia, Indonesia, Lao PDR, Malaysia, Myanmar, Philippines, Singapore, Thailand, Timor-Leste, and Vietnam. Such a design allows us to compare the data reporting practices between different countries through time as the pandemic progresses.^14^

### Data sources and compilation

We focused on reporting mechanisms of individual level COVID-19 data from the aforementioned 11 countries in Southeast Asia. The region is characterized by archipelagos and comprises more than 8·0% of the world’s population. During the first wave of the pandemic, these 11 countries contributed about 1·3% of the cases to the global count of more than 2·3 million cases on April 20.

The Open COVID-19 Data Curation Group’s centralised repository of individual-level information on patients with laboratory-confirmed COVID-19 collected data on the following variables deemed essential in monitoring pandemics: (a) Key dates, which include the date of travel, date of onset of symptoms, date of confirmation of infection, date of admission to hospital, and date of outcome; (b) Demographic information inclduing the age, sex, and occupation of cases; (c) Geographic information on domicile and travel history at the highest resolution available down to the district level; (d) Any additional information such as symptoms and ‘contact tracing data’ (i.e., a record of exposure to infected individuals).^10^ Information about the occupation of confirmed cases was also included where available. The collection of data on these variables mirrors the minimum data to be collected for a line list of pandemic influenza cases obtained from surveillance systems, as suggested by the WHO.^15^ Other sources, such as the interactive dashboard by Johns Hopkins University,^9^ do not provide detailed individual-level information and hence were not used in this study.

We reviewed the data of the Open COVID-19 Data Working Group’s centralised repository and other relevant data sources. These were primarily websites of the various governments and ministries of health in Southeast Asia, including but not limited to their COVID-19 trackers, dashboards and case bulletins. In addition, we reviewed data from news agencies, pre-prints and peer-reviewed research articles that contained information on COVID-19 cases in the country. We reviewed all possible publicly available data sources from the date when the first confirmed case was reported in the country, and up to April 20. We documented how key information was reported and how it changed through time.

### Shifts in reporting of epidemiological data during the first wave

We documented trends and changes in how key epidemiological variables were reported by 11 Southeast Asian countries throughout the study period from January 23 to April 20. Time periods were defined by specific milestones in each country’s data reporting. The first time period or ‘first reporting of cases’ for all countries was the date at which the country reported its first COVID-19 case. Meanwhile, the ‘first change in reporting’ was the time when the information format was changed from the first report based on available data during the study period. Any further changes in the level of detail, also referred in this paper as granularity for geographic data and precision for both demographic and temporal data, in the reporting of any of the epidemiological variables were considered as a ‘change in reporting’ and were noted as a subsequent time period (Table 1). The ‘last observed change in reporting’ was the last documented change up to April 20. We also noted the number of cases in each timepoint. In this paper, we only present results on the ‘first reporting of cases’ (T0), ‘first observed change in reporting’ (T1), and ‘last observed change in reporting’ (T2).

**Table 1.** Variables included in the study and the definitions of ‘change in reporting’.

| Variable | Definition and possible responses | Change in reporting |
| --- | --- | --- |
| Date of reporting | Date at which first case of COVID-19 is reported or date at which there is change of reporting in any of the variables | Not applicable |
| Cases | Number of cases as of the ‘Date of reporting’ | Not applicable |
| Data aggregation | Type of data aggregation used. “Individual” denotes that information is reported for each individual case, “Aggregated” if otherwise. | Country no longer reports individual-level data or vice versa |
| *Demographic* |  |  |
| Age | Reporting of age of COVID-19 cases. “Exact” if age in years is reported for each individual case. “Age bracket” if the country reports age ranges. “None” if not reported. | Country’s level of reporting shifts between “Exact”, “Age bracket”, or “None” |
| Sex | Reporting of sex of COVID-19 cases. “With” if the country reports the sex of their individual confirmed cases. “Without” if not reported. | Country no longer reports data on sex after reporting it previously, or vice versa. |
| Occupation | Reporting of occupation of COVID-19 cases. “With” if the country reports the occupation of individual confirmed cases. “Without” if not reported. | Country no longer reports occupation data after reporting it previously or vice versa |
| *Geographic* |  |  |
| Domicile | Reporting of domicile of COVID-19 cases. “Precise location” if street-level addresses or exact location information are available. “City” if city addresses are available. “Province” if province/state-level addresses are available. “Country” if only country-level addresses are available. “None” if not reported. | Country changes the level of precision at which it reports its domicile data (e.g., precise location to province) |
| Travel history location | Reporting of international travel history of COVID-19 cases. “Precise location” if street-level addresses or exact location information are available. “City” if city addresses are available. “Province” if province/state-level addresses are available. “Country” if only country-level addresses are available. “None” if not reported. | Country changes the level of precision at which it reports its domicile data (e.g., precise location to province) |
| *Temporal* |  |  |
| Date of travel | Reporting of dates of international travel of COVID-19 cases. “Day” if the country reports the exact date of travel. “Month” if the country only reports the month of travel. “None” if it is not reported. | Country’s level of reporting shifts between “Day, “Month”, and “None” |
| Date of symptom onset | Reporting of date of symptom onset of COVID-19 cases. “Day” if the country reports the exact date of travel. “Month” if the country only reports the month of travel. “None” if it is not reported. | Country’s level of reporting shifts between “Day, “Month”, and “None” |
| Date of confirmation | Reporting of date of laboratory confirmation of COVID-19 cases. “Day” if the country reports the exact date of confirmation. “Month” if the country only reports the month of confirmation. “None” if it is not reported. | Country’s level of reporting shifts between “Day, “Month”, and “None” |
| Date of hospital admission | Reporting of date of hospital admission of COVID-19 cases. “Day” if the country reports the exact date of hospital admission. “Month” if the country only reports the month of hospital admission. “None” if it is not reported. | Country’s level of reporting shifts between “Day, “Month”, and “None” |
| Date of discharge, recovery, or death | Reporting of date of hospital discharge or recovery or death of COVID-19 cases. “Day” if the country reports the exact date of hospital discharge or death. “Month” if the country only reports the month of hospital discharge or death. “None” if it is not reported. | Country’s level of reporting shifts between “Day, “Month”, and “None” |
| *Other epidemiological variables* |  |  |
| Travel history | Reporting of international travel history of COVID-19 cases. “With” if the country reports the occupation of individual confirmed cases. “Without” if not reported. | Country no longer reports travel history data after reporting it previously or vice versa |
| Symptoms | Reporting of any symptoms of COVID-19 cases. “With” if the country reports the occupation of individual confirmed cases. “Without” if not reported. | Country no longer reports symptom data after reporting it previously or vice versa |
| Comorbidities | Reporting of any chronic disease or comorbidity of COVID-19 cases. “With” if the country reports the occupation of | Country no longer reports comorbidities after reporting it previously or vice versa |
|  | individual confirmed cases. “Without” if not reported. |  |
| Cluster | Reporting of disease clusters or known contacts of COVID-19 cases. “With” if the country reports the occupation of individual confirmed cases. “Without” if not reported. | Country no longer reports cluster data after reporting it previously or vice versa |
| Outcome | Reporting of hospital discharge or recovery or death of COVID-19 cases. “With” if the country reports the occupation of individual confirmed cases. “Without” if not reported. | Country no longer reports outcome data after reporting it previously or vice versa |

### Differences in the granularity and precision of reporting across countries

We explored the differences in reporting of demographic, geographic, and temporal data across countries at three key timepoints: at the time they first reported cases (T0), at the time when the reporting first changed (T1), and at the last observed change in reporting (T2). Any change in the level of granularity or precision in reporting is noted. We present these differences for each epidemiological variable classified into: (a) demographic data; (b) geographic data; and (c) temporal data. Data for other epidemiological variables are presented in the Supplementary Appendix.

### Accessibility of COVID-19 data in the region

We described the accessibility of COVID-19 data in the region by documenting and providing a list of sources for each country in different formats, including: government trackers and dashboards that report close to real-time data, downloadable PDF reports on cases, downloadable CSV files for cases, and Github repository for cases. In addition, we noted the frequency of data updates by government and public health agencies in the country.

## Results

### Shifts in reporting of epidemiological data during the first wave

The first Southeast Asian country to report a COVID-19 case was Thailand on January 23. Singapore, Malaysia, Cambodia, Vietnam and the Philippines subsequently reported cases on or before the WHO declared COVID-19 a Public Health Emergency of International Concern (PHEIC) on January 30. Indonesia, Brunei, Timor-Leste, Myanmar and Lao PDR reported their first cases of COVID-19 in March (Figure 1).

**Figure 1.**
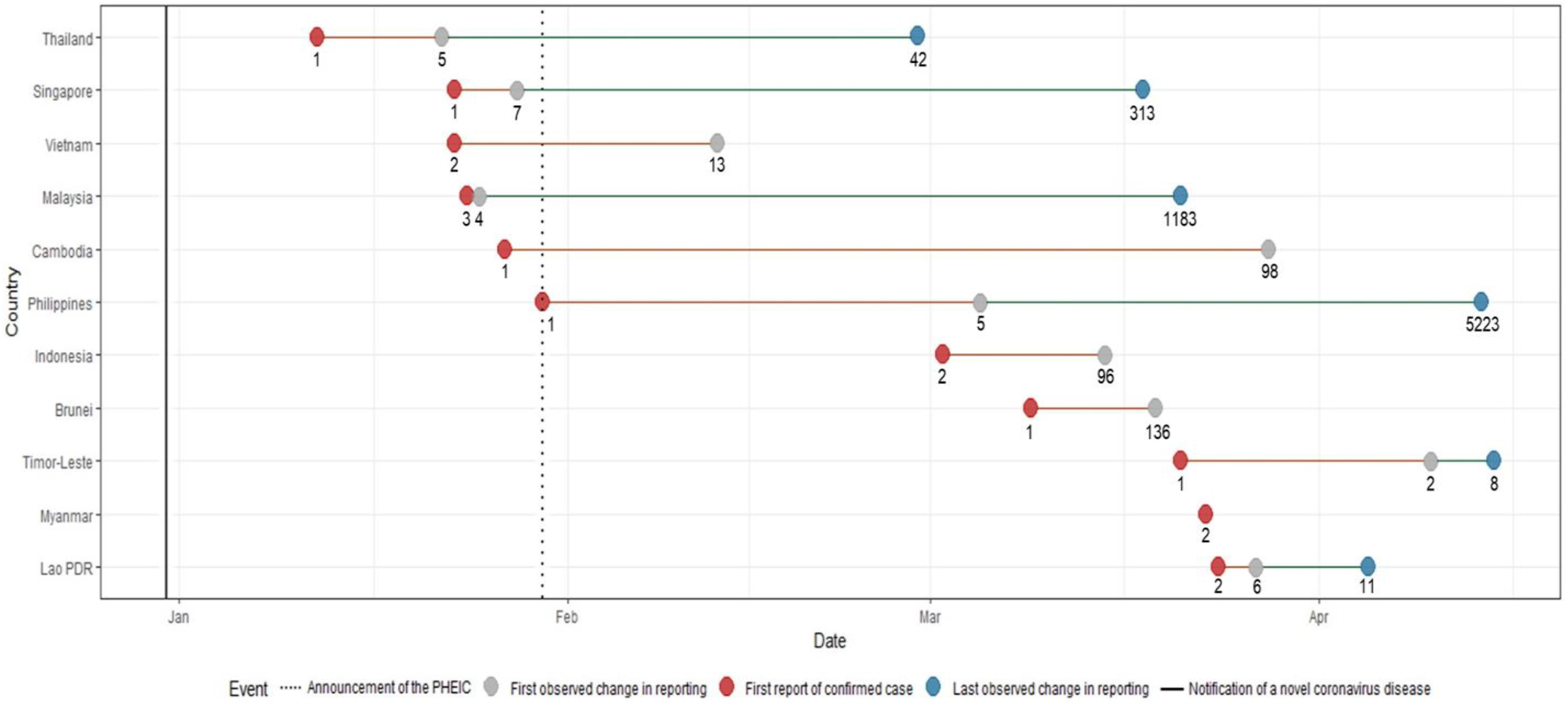
Timeline of key events with corresponding number of cases during the first wave of the COVID-19 pandemic. The notification of a novel coronavirus disease in late December and the announcement of the PHEIC are denoted by lines. Shifts in reporting are defined by a change in level of detail and precision in any of the epidemiological variables. Each key reporting shift is denoted by a colored circle.

Malaysia had the shortest time between reporting of the first case and first change in reporting of epidemiological data. Only a day after their first reported case, more detailed reports on the occurrence of symptoms, and dates of symptom onset and hospitalisation were provided. Similar improvements in terms of the level of granularity and precision in reporting data were also noted for the following countries: Philippines eventually reported comorbidities for some patients, Singapore and Vietnam eventually reported data on occupation, and Timor-Leste eventually reported travel history data. As case numbers increased, several countries provided less detailed information. By March 15, when 96 cases had been identified, Indonesia ceased reporting individual-level data and switched to aggregate data (i.e., number of cases per day). Timor-Leste followed by April 15, when it had 8 recorded cases. The first and the last changes in reporting were the same for Indonesia and Brunei, while Myanmar was the only country that consistently reported individual-level COVID-19 epidemiologic data since reporting its first two cases on March 23 until April 20.

### Differences in the granularity and precision of reporting across countries

There were minimal changes in the reporting of demographic data among countries. The majority of countries reported age and sex except for Timor-Leste, and only Indonesia shifted from a more precise reporting of age and sex to less detailed reporting (Figures 2a and 2b). We observed more changes in the reporting of occupation (Figure 2c); Indonesia only provided occupation data at the time of reporting of first cases, while Singapore and Vietnam included data on occupation of COVID-19 patients at later timepoints.

**Figure 2.**
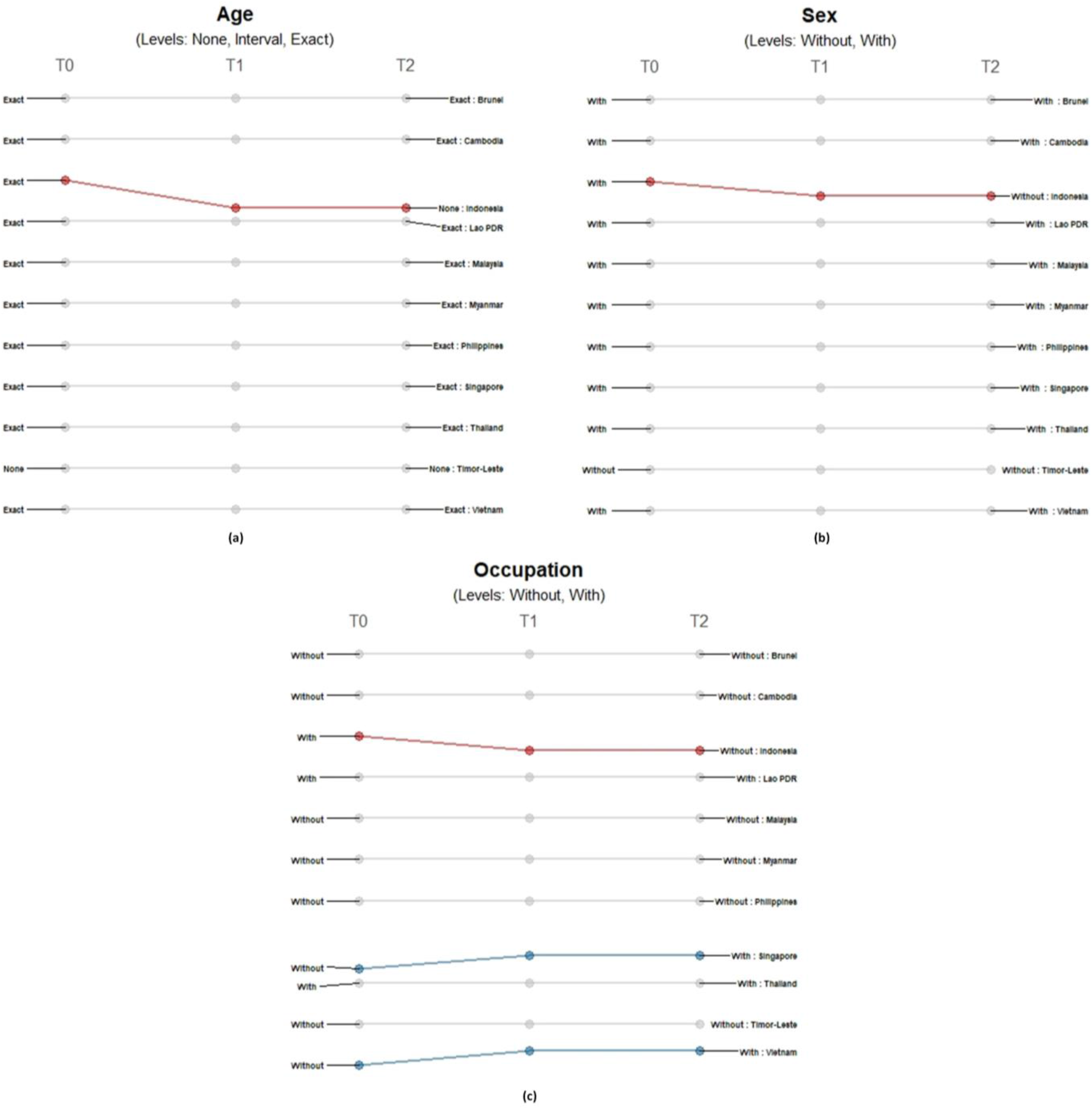
Differences in the level of precision in reporting demographic data: (a) age, (b) sex, and (c) occupation over three timepoints. Only those countries with changes in the level of detail and precision of reporting are highlighted. Each country may shift reporting at any timepoint: at the first reporting of cases’ (T0), ‘first observed change in reporting’ (T1), and ‘last observed change in reporting’ (T2). Each country may report less precise data indicated by a decreasing slope (red) or more precise data indicated by an increasing slope (blue) consistently over time. Reporting may not be consistent across timepoints with shifts between different levels of precision (yellow) or reporting may not have changed at all during the study period (grey). The levels of precision are indicated for each epidemiological variable. Age has three levels while both sex and occupation are binary variables.

Location information on domicile and travel history differed across countries and timepoints. While all 11 countries provided domicile information (Figure 3a), only Singapore provided precise-level addresses. Both Indonesia and Malaysia initially provided city-level information and shifted to less granular reporting. For Indonesia, province-level data was being reported by March 15 when it reached 96 cases. Meanwhile, Malaysia started reporting province-level data on March 21 when it reached 1183 confirmed cases. On the other hand, the information coming from some countries initially presented less granularity or lower geographic resolution: Lao PDR initially reported country-level information, Thailand initially reported province-level addresses and Vietnam initially reported city-level addresses; eventually all three countries reported precise address data. There were less differences observed for travel location data reporting across countries, but also more shifts observed across time (Figure 3b). Most (8 of 11) provided city-level information of the travel history; only Myanmar provided country-level information, while both Indonesia and Timor-Leste provided no information at the time of reporting their first cases. Only Timor-Leste shifted to a more precise level of reporting over time, while Brunei, Cambodia, Malaysia, Philippines, Singapore and Thailand reported less granular data. Lao PDR shifted reporting travel histories from city-level information when it reported its first two cases to no information being shared when it had six confirmed cases, and then to country-level travel history data when it had reported 11 cases.

**Figure 3.**
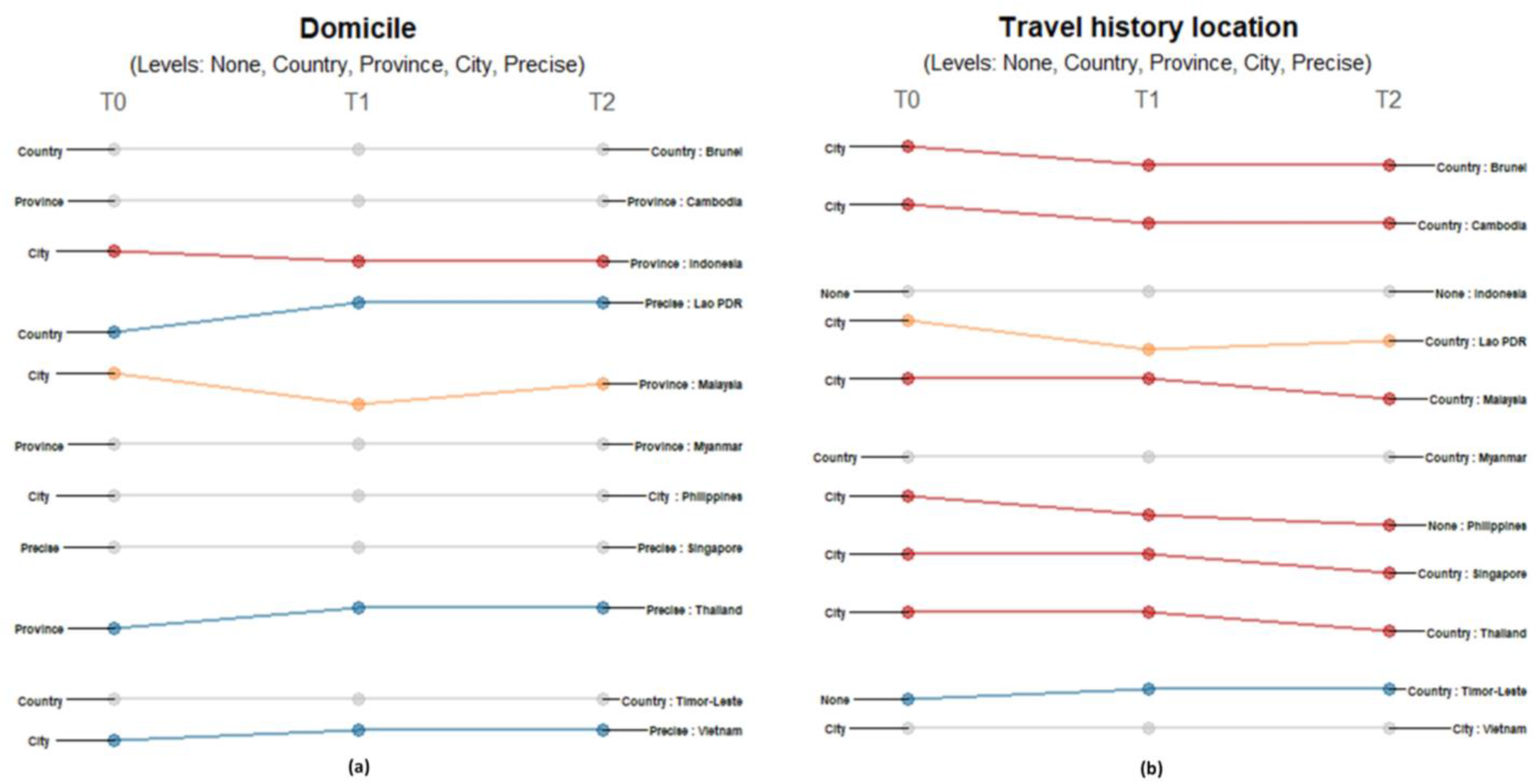
Differences in the level of granularity in reporting geographic data: (a) domicile, and (b) travel history location over three timepoints. Only those countries with changes in the level of granularity or geographic resolution of reporting are highlighted. Each country may shift reporting at any timepoint: at the first reporting of cases’ (T0), ‘first observed change in reporting’ (T1), and ‘last observed change in reporting’ (T2). Each country may report less granular data indicated by a decreasing slope (red) or more granular data indicated by an increasing slope (blue) consistently over time. Reporting may not be consistent across timepoints with shifts between different levels of granularity (yellow) or reporting may not have changed at all during the study period (grey). The levels of granularity are indicated for each epidemiological variable. All geographic data have five levels of granularity/geographic resolution: none, country, province, city, and precise.

For all temporal variables, countries reported either precise dates or no dates at all. At the start of each country’s first case, the majority of countries provided travel history dates except for Brunei, Indonesia, and Timor-Leste (Figure 4a). Only Brunei shifted to reporting dates for the succeeding timepoints while Malaysia, Philippines, and Singapore stopped reporting dates as cases increased. Lao PDR repeatedly shifted between reporting travel dates and excluding this information. The precision of reporting symptom onset dates also varied across countries and timepoints (Figure 4b). Cambodia, Indonesia and Timor-Leste never reported such information, while Brunei and Vietnam consistently reported specific dates when symptoms presented. Malaysia provided day information in the succeeding timepoints while Myanmar, Philippines, Singapore, and Thailand eventually stopped reporting the date of symptom onset. Lao PDR repeatedly shifted between reporting of dates to no reporting. Both date of confirmation and date of outcome showed consistent reporting in all countries except Thailand, which stopped reporting the date of confirmation when it reported 42 cases (Figure 4c). Most countries reported the date of admission except for Brunei, Cambodia, Indonesia, Malaysia, and Timor-Leste (Figure 4d). Only Thailand had a shift in reporting dates of discharge, recovery, or death - reporting this information only in late February when it had 42 cases (Figure 4e). Malaysia provided this information in the succeeding timepoints while Philippines, Singapore, and Thailand eventually stopped reporting the date of symptom onset. Lao PDR repeatedly shifted between reporting of dates to providing no such information.

**Figure 4.**
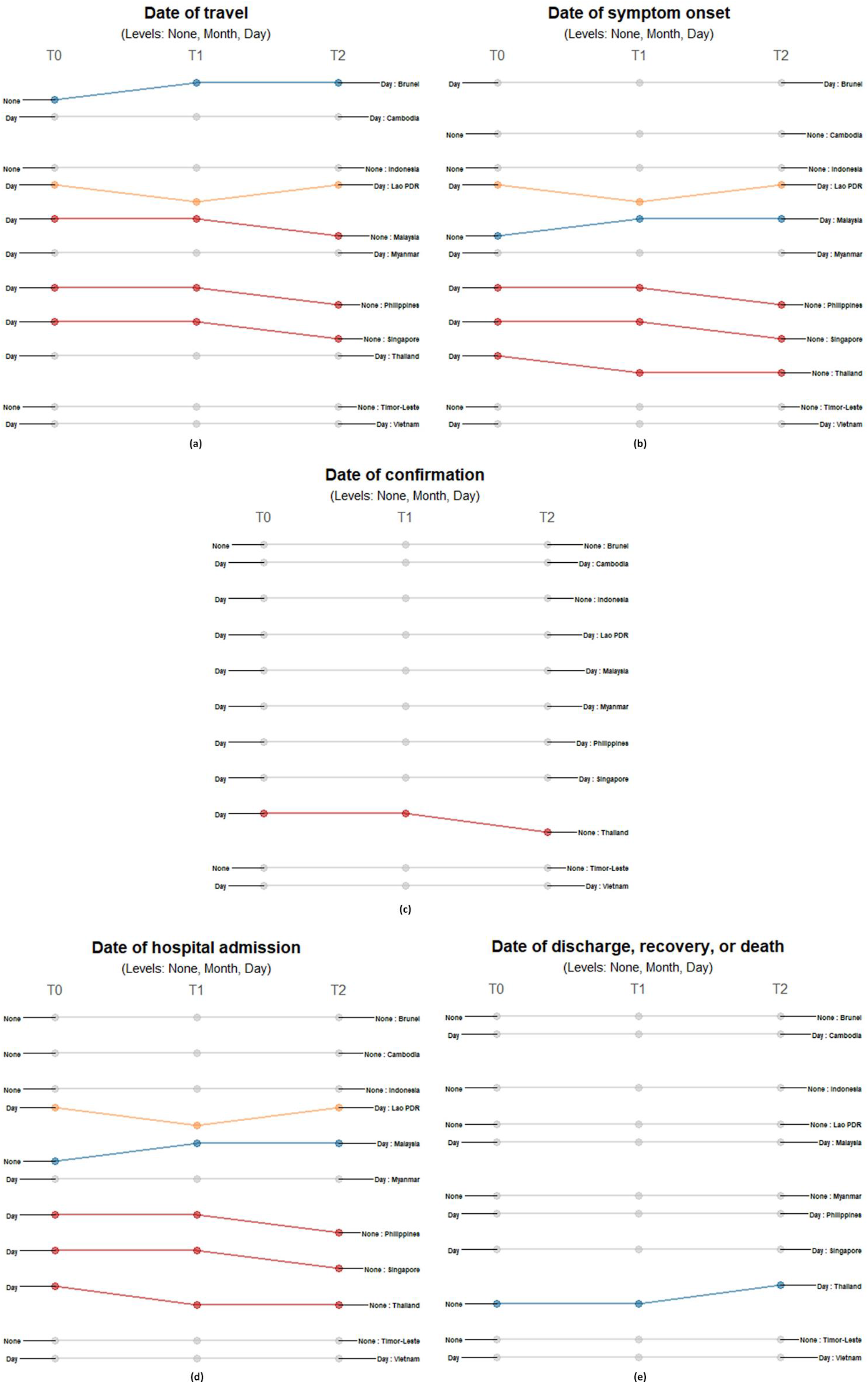
Differences in the level of precision in reporting temporal data: (a) date of travel, (b) date of symptom onset, (c) date of confirmation, (d) date of hospital admission, and (e) date of outcome over three timepoints. Only those countries with changes in the level of detail and precision of reporting are highlighted. Each country may shift reporting at any timepoint: at the first reporting of cases’ (T0), ‘first observed change in reporting’ (T1), and ‘last observed change in reporting’ (T2). Each country may report less precise data indicated by a decreasing slope (red) or more precise data indicated by an increasing slope (blue) consistently over time. Reporting may not be consistent across timepoints with shifts between different levels of precision (yellow) or reporting may not have changed at all during the study period (grey). The levels of precision are indicated for each epidemiological variable. All date variables have three levels of precision: none, month, and day.

### Accessibility of COVID-19 data in the region

Most of the governments in Southeast Asia instituted infrastructures and guidelines for the dissemination of COVID-19 data, and provided free access to the public. Brunei’s COVID-19 data are publicly available but the individual requesting the information needs to provide passport information and contact number to gain access to the database. During the study period, all countries had government trackers and dashboards except for Timor-Leste (Table 2). Cambodia, Indonesia, Malaysia, Philippines, Singapore, Thailand, and Vietnam provide daily updates on their COVID-19 data. For the remaining five countries, the frequency of updating is unclear/irregular. Brunei, Philippines, and Singapore use PDFs to disseminate select patient-level data, and only Thailand provides downloadable individual-level CSV files.

**Table 2.** Availability of government dashboards and trackers for COVID-19, frequency of updates, and raw data sharing formats used in countries in Southeast Asia as of April 20.

| Country | Availability of government tracker/ dashboard | Frequency of update | Raw data sharing formats ever used |  |  |  |
| --- | --- | --- | --- | --- | --- | --- |
|  |  |  | HTML | CSV | PDF | Other |
| Brunei | Available<sup>16</sup> | Unclear | Yes | No | Yes | None |
| Cambodia | Available<sup>17</sup> | Daily | No | No | No | None |
| Indonesia | Available<sup>18-20</sup> | Daily | No | No | No | None |
| Lao PDR | Available<sup>21</sup> | Unclear | Yes<sup>22</sup> | No | No | None |
| Malaysia | Available<sup>23</sup> | Daily | Yes<sup>24</sup> | No | No | None |
| Philippines | Available<sup>25</sup> | Daily | Yes<sup>25</sup> | No | Yes<sup>26</sup> | Google Sheets<sup>27</sup> |
| Singapore | Available<sup>28</sup> | Daily | Yes<sup>29</sup> | No | Yes<sup>29</sup> | None |
| Thailand | Available<sup>30</sup> | Daily | No | Yes<sup>31</sup> | No | None |
| Timor-Leste | Not available | Unclear | No | No | No | None |
| Vietnam | Available<sup>32</sup> | Daily | Yes<sup>32</sup> | No | No | None |

## Discussion

Responding to calls for data sharing and transparency, most governments in Southeast Asia established publicly available sources of COVID-19 individual-level information. This commitment to data sharing and reporting allowed the comparison of the different data reporting practices of the countries in the region. We found that countries in Southeast Asia have different reporting practices since the start of the pandemic and during the first months of its progression. Overall, reporting of epidemiological data in Southeast Asia is precise and detailed. Many variables were consistently maintained throughout the initial outbreak period, but those with changes in reporting started early with case counts as low as four to as high as 136. There was little to no change in reporting of demographic data while changes in reporting of geographic and temporal variables were frequent and unpredictable as the pandemic progressed. Further, we find that changes in the level of precision in reporting does not only depend on case numbers, but also on the policies and interventions implemented. Comparisons across countries for different epidemiologic variables showed that national governments may shift to a less or more precise reporting of data as dictated by the burden of COVID-19 in the communities and/or their national response. As an example, Indonesia started reporting aggregate data less than two weeks after their first case was reported. Their government did not implement a nationwide lockdown, but rather focused on scaling up capacity, treatment of patients and supporting economic recovery. Conversely, Lao PDR, Thailand and Vietnam reported more precise demographic and geographic data at the end of the study period compared to how they reported their first cases. The national governments of these countries established mechanisms to quickly identify and isolate cases and their contacts requiring detailed contact tracing data. Our findings also show that most countries reported more precise information towards the end of the study period, but some variables such as travel history location were reported with less detail compared to the increased precision for domicile data. These trends in travel history data highlight the shift in priorities of the governments in the region towards managing local transmission. Southeast Asian countries implemented travel restrictions early, therefore having fewer imported cases and less need for precise travel history data.^33^

Data on dates of symptom onset, confirmation, admission, and death or discharge are important in estimating disease burden and forecasting health service needs. Dates of confirmation and death or discharge were reported consistently by most countries. This reflects the effective system of governments to register all confirmed patients in their database upon entry and exit in the healthcare system. However, we found that dates of symptom onset and hospital admission were no longer reported at the end of the observation period. The reporting of less precise dates could be attributed to the increasing incidence of COVID-19, which could have overwhelmed data reporting mechanisms of the countries, particularly because individual patient follow-up requires symptom onset dates to be accurately logged. Governments thus need to establish systems that allow accurate and fast reporting of detailed temporal data. Lack of precision could adversely affect the quality of mathematical models and other analyses, which are used to forecast demand for health services and make decisions. This consequently impacts the responses to COVID-19 at a national and subnational level, which is of greater concern among low- and middle-countries (LMICs) that have already fragile health systems. Our findings provide insights on how different health systems respond to the pandemic. Consequently, these could be used to guide how publicly available data is analysed, used, and interpreted.

Most countries reported COVID-19 data daily, with unclear reporting frequencies only being observed for Brunei, Lao PDR, and Timor-Leste. These countries do not report new cases every day because of the low number of new daily cases leading to days where no additional cases are confirmed. As they only provide updates on days when new COVID-19 cases are confirmed, their frequency of providing data updates on COVID-19 is thus irregular. Countries primarily reported individual-level data in either HTML and PDF formats, which necessitates scraping and extraction before such data could be used in analyses. During the study period, only Thailand provided a downloadable CSV format of their data. Ready-to-use data formats are important as these allow the public and scientific community to rapidly view and analyse country-specific information.

An important limitation of this study is the absence of an assessment on data quality. This evaluation was not carried out because of the fast progression of the pandemic with corresponding rapid changes in data reporting. The lack of an up-to-date and complete line list also prevents a thorough assessment of data quality. Lastly and most importantly, an evaluation of data quality also requires the consideration of other indicators such as flexibility, representativeness, data security and system stability to provide a more accurate picture of health systems and disease surveillance systems.^7^ These information are not readily available and require more resources to be collected. Despite such caveats, however, this study is the first to systematically describe and compare reporting of important epidemiological data for COVID-19 across countries during the first wave. Our findings will allow researchers to conduct more nuanced analyses using epidemiological data of COVID-19.

In conclusion, reporting systems in the region have been quickly established and countries provided detailed individual-level data during the first wave. This pandemic highlights the critical role of timely, accurate, and precise data sharing during outbreaks of global scale. Some concerns regarding data sharing remain, such as data privacy and public criticisms.^3,4^ Given that sharing of data is needed for evidence-informed policies and interventions, maintaining and strengthening data reporting systems should still be a priority of countries.^34-36^ For the purposes of surveillance on emerging infectious diseases, we recommend that governments coordinate data collection and reporting so that data are as comparable as possible between countries. Countries may also benefit from reporting data in a fully open access format that is readily available and in machine-readable formats to accommodate new epidemics and context-specific information. Hopefully, more governments will come to share precise data to allow more nuanced analyses. This will provide an opportunity to better understand the disease and how best to respond to the pandemic.

## Data Availability

The data that support the findings of this study are openly available in GitHub at https://github.com/beoutbreakprepared/nCoV2019.

## Author contributions

All authors contributed to the study concept and design, data compilation, analysis, and interpretation. AMLA and VCFP wrote the initial draft of the manuscript with inputs from BG and TR. All authors critically revised the report and approved the final version for submission.

## Data sharing

All data and code are available here: https://github.com/beoutbreakprepared/nCoV2019

## Acknowledgements

We thank Moritz U.G. Kraemer and the Open COVID-19 Data Working Group for their support and insights. The full list of curators and contributors making up the Open COVID-19 Data Working Group is provided at: https://github.com/beoutbreakprepared/nCoV2019. This work was supported through an Engineering and Physical Sciences Research Council (EPSRC) (https://epsrc.ukri.org/) Systems Biology studentship award (EP/G03706X/1) to TR. This project was also supported in part by the Oxford Martin School.

## References

1. Dye C, Bartolomeos K, Moorthy V, Kieny MP. Data sharing in public health emergencies: a call to researchers. Bull World Health Organ. 2016 Mar 1;94(3):158–158.

2. Pearce N, Vandenbroucke JP, VanderWeele TJ, Greenland S. Accurate Statistics on COVID-19 Are Essential for Policy Guidance and Decisions. Am J Public Health. 2020 Apr 23;e1–3.

3. Littler K, Boon W-M, Carson G, Depoortere E, Mathewson S, Mietchen D, et al. Progress in promoting data sharing in public health emergencies. Bull World Health Organ. 2017 Apr 1;95(4):243–243.

4. Chretien J-P, Rivers CM, Johansson MA. Make Data Sharing Routine to Prepare for Public Health Emergencies. PLOS Medicine. 2016 Aug 16;13(8):e1002109.

5. Wilkinson MD, Dumontier M, Aalbersberg IjJ, Appleton G, Axton M, Baak A, et al. The FAIR Guiding Principles for scientific data management and stewardship. Scientific Data. 2016 Mar 15;3(1):1–9.

6. GloPID-R. Data Sharing Working GroupGroup. Principles for Data Sharing in Public Health Emergencies [Internet]. 2017 [cited 2020 May 7]. Available from: https://wellcome.figshare.com/articles/Principles_for_Data_Sharing_in_Public_Health_Emergencies/4733590.

7. Lawpoolsri S, Kaewkungwal J, Khamsiriwatchara A, Sovann L, Sreng B, Phommasack B, et al. Data quality and timeliness of outbreak reporting system among countries in Greater Mekong subregion: Challenges for international data sharing. PLOS Neglected Tropical Diseases. 2018 Apr 25;12(4):e0006425.

8. Moorthy V, Henao Restrepo AM, Preziosi M-P, Swaminathan S. Data sharing for novel coronavirus (COVID-19). Bull World Health Organ. 2020 Mar 1;98(3):150.

9. Dong E, Du H, Gardner L. An interactive web-based dashboard to track COVID-19 in real time. The Lancet Infectious Diseases. 2020 May 1;20(5):533–4.

10. Xu B, Kraemer MUG, Xu B, Gutierrez B, Mekaru S, Sewalk K, et al. Open access epidemiological data from the COVID-19 outbreak. The Lancet Infectious Diseases. 2020 May 1;20(5):534.

11. Sun K, Chen J, Viboud C. Early epidemiological analysis of the coronavirus disease 2019 outbreak based on crowdsourced data: a population-level observational study. The Lancet Digital Health. 2020 Apr;2(4):e201–8.

12. Wang LL, Lo K, Chandrasekhar Y, Reas R, Yang J, Burdick D, et al. CORD-19: The COVID-19 Open Research Dataset. ArXiv200410706 Cs [Internet]. 2020 Jul 10 [cited 2020 Oct 21]; Available from: http://arxiv.org/abs/2004.10706.

13. Tsang TK, Wu P, Lin Y, Lau EHY, Leung GM, Cowling BJ. Effect of changing case definitions for COVID-19 on the epidemic curve and transmission parameters in mainland China: a modelling study. The Lancet Public Health. 2020 May 1;5(5):e289–96.

14. Rothman K, Greenland S, Lash T. Modern Epidemiology. >Third Edition. Philadelphia, PA: Lippincott Williams and Wilkins; 2008.

15. World Health Organization. WHO guidance for surveillance during an influenza pandemic [Internet]. 2017 [cited 2020 May 16]. Available from: https://www.who.int/influenza/preparedness/pandemic/WHO_Guidance_for_surveillance_during_an_influenza_pandemic_082017.pdf.

16. Ministry of Health (Brunei). Welcome to HealthInfo [Internet]. 2020 [cited 2020 Jun 17]. Available from: https://www.healthinfo.gov.bn/smartguide/user/login?callback=https%3A%2F%2Fwww.healthinfo.gov.bn%2F%23%2Fhome.

17. Ministry of Health (Cambodia). 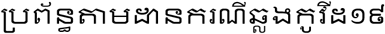 [Internet]. 2020 [cited 2020 Jun 17]. Available from: https://covid19-map.cdcmoh.gov.kh/?fbclid=IwAR29DjPwYCLbkqtreXbXnRgsAVjOqEAA0HnX96NQnLLdeT4XKep_PDwH-3U.

18. Ministry of Health (Indonesia). Home » Info Infeksi Emerging Kementerian Kesehatan RI [Internet]. Info Infeksi Emerging Kementerian Kesehatan RI. [cited 2020 Jun 17]. Available from: https://covid19.kemkes.go.id/.

19. Gugus Tugas Percepatan Penanganan COVID-19. Home Task Force for the Acceleration of Handling COVID-19 [Internet]. covid19.go.id. 2020 [cited 2020 Jun 17]. Available from: https://covid19.go.id/.

20. Ariansyah A. Badan Nasional Penanggulangan Bencana [Internet]. BNPB. 2020 [cited 2020 Jun 17]. Available from: https://bnpb.go.id.

21. Ministry of Health (Laos). 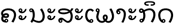 COVID-19 [Internet]. [cited 2020 Jun 17]. Available from: https://www.covid19.gov.la/index.php.

22. Ministry of Health (Laos). 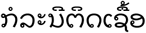[Internet]. 2020 [cited 2020 Jun 17]. Available from: https://www.covid19.gov.la/index.php?r=site%2Fdetail&id=329.

23. Evozi. COVID-19 | Malaysia Outbreak Monitor | Live Updates [Internet]. COVID-19 | Malaysia Outbreak Monitor | Live Updates. 2020 [cited 2020 Jun 17]. Available from: https://www.outbreak.my.

24. Evozi. Malaysia - Statistics | COVID-19 | Malaysia Outbreak Monitor | Live Updates [Internet]. Malaysia - Statistics | COVID-19 | Malaysia Outbreak Monitor | Live Updates. 2020 [cited 2020 Jun 17]. Available from: https://www.outbreak.my/stats.

25. Department of Health (Philippines). COVID-19 Tracker Philippines [Internet]. 2020 [cited 2020 Jun 17]. Available from: https://ncovtracker.doh.gov.ph/.

26. Department of Health (Philippines). Updates of Novel Coronavirus Disease (COVID-19) [Internet]. [cited 2020 Mar 19]. Available from: https://www.doh.gov.ph/2019-nCov.

27. Department of Health (Philippines). DOH COVID-19 DataDrop - Google Drive [Internet]. 2020 [cited 2020 Jun 10]. Available from: https://drive.google.com/drive/folders/10VkiUA8x7TS2jkibhSZK1gmWxFM-EoZP.

28. Ministry of Health (Singapore). COVID-19: Cases in Singapore [Internet]. 2020 [cited 2020 Jun 17]. Available from: http://www.gov.sg/article/covid-19-cases-in-singapore.

29. Ministry of Health (Singapore). MOH | News Highlights [Internet]. 2020 [cited 2020 Jun 17]. Available from: https://www.moh.gov.sg/news-highlights/details/797-more-cases-discharged-151-new-cases-of-covid-19-infection-confirmed.

30. Digital Government Development Agency. 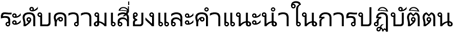 COVID19 (Update 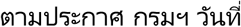 7/4/2020) - Open Government Data of Thailand [Internet]. 2020 [cited 2020 Jun 17]. Available from: https://data.go.th/en/dataset/covid19.

31. Digital Government Development Agency (Thailand).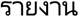 COVID-19 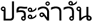 – Open Government Data of Thailand [Internet]. 2020 [cited 2020 Jun 17]. Available from: https://data.go.th/en/dataset/covid-19-daily.

32. Ministry of Health (Vietnam). TRANG TIN VỀ DỊCH BỆNH VIÊM ĐƯỜNG HÔ HẤP CẤP COVID-19 - Bộ Y tế - Trang tin về dịch bệnh viêm đẰờng hô hấp cấp COVID-19 [Internet]. 2020 [cited 2020 Jun 17]. Available from: https://ncov.moh.gov.vn/.

33. World Health Organization. Pandemic Influenza Preparedness and Response: A WHO Guidance Document [Internet]. Pandemic Influenza Preparedness and Response: A WHO Guidance Document. World Health Organization; 2009 [cited 2020 Aug 6]. Available from: https://www.ncbi.nlm.nih.gov/books/NBK143061/.

34. Goldacre B, Harrison S, Mahtani K, Heneghan C. WHO consultation on Data and Results Sharing During Public Health Emergencies [Internet]. Center for Evidence-Based Medicine, University of Oxford; 2015 [cited 2020 Jun 18]. Available from: https://www.who.int/medicines/ebola-treatment/background_briefing_on_data_results_sharing_during_phes.pdf?ua=1.

35. Aljunid SM, Srithamrongsawat S, Chen W, Bae SJ, Pwu R-F, Ikeda S, et al. Health-Care Data Collecting, Sharing, and Using in Thailand, China Mainland, South Korea, Taiwan, Japan, and Malaysia. Value in Health. 2012 Jan 1;15(1, Supplement):S132–8.

36. Gewin V. Six tips for data sharing in the age of the coronavirus. Nature [Internet]. 2020 May 19 [cited 2020 Jun 18]; Available from: https://www.nature.com/articles/d41586-020-01516-0.

