## Supplementary Appendix for "Tracking changes in reporting of epidemiological data during the COVID-19 pandemic in Southeast Asia: an observational study during the first wave"

**Supplementary Material**

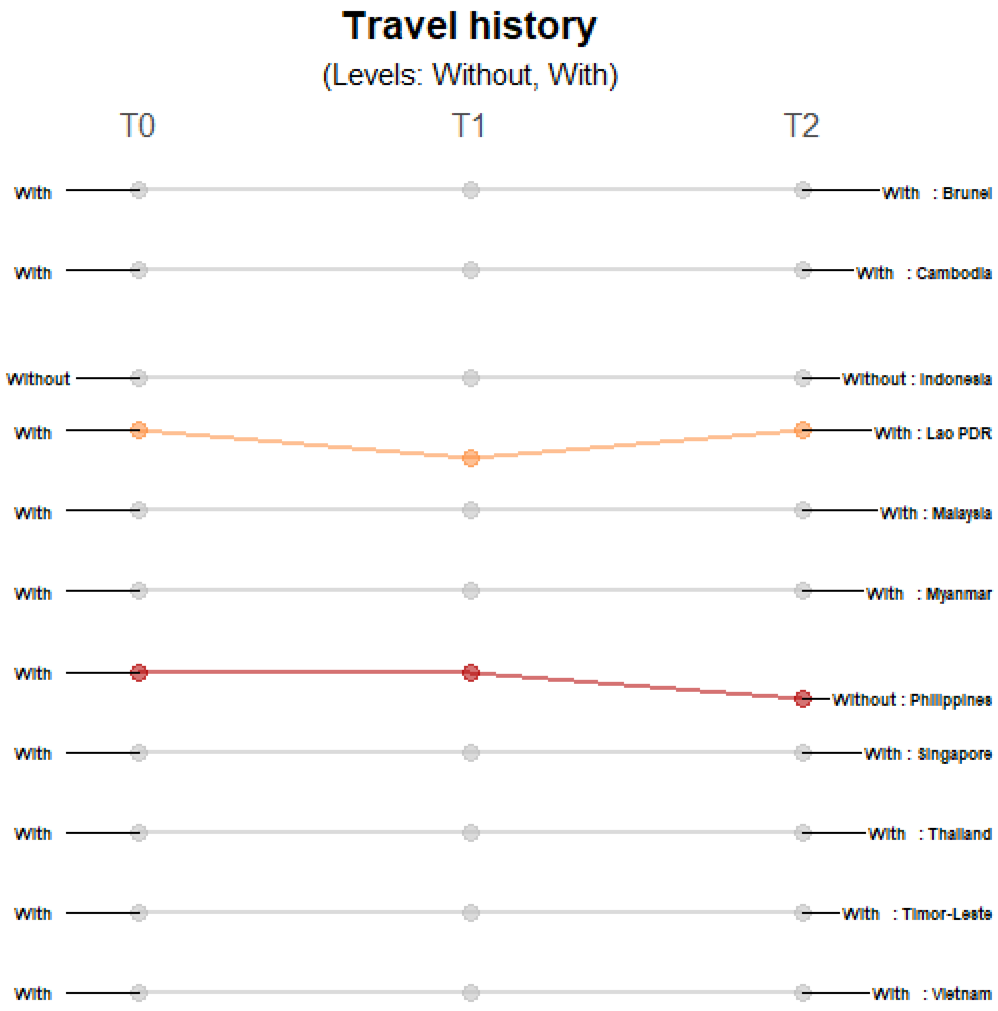


**Supplementary Figure 1. Differences in the level of precision in reporting travel history**. Only those countries with changes in the level of detail and precision of reporting are highlighted. Each country may shift reporting at any timepoint: at the first reporting of cases' (T0), 'first observed change in reporting' (T1), and 'last observed change in reporting' (T2). Each country may report less precise data indicated by a decreasing slope (red) or more precise data indicated by an increasing slope (blue) consistently over time. Reporting may not be consistent across timepoints with shifts between different levels of precision (yellow) or reporting may not have changed at all during the study period (grey). The levels of precision are indicated for this epidemiological variable. This variable has two levels: without and with.

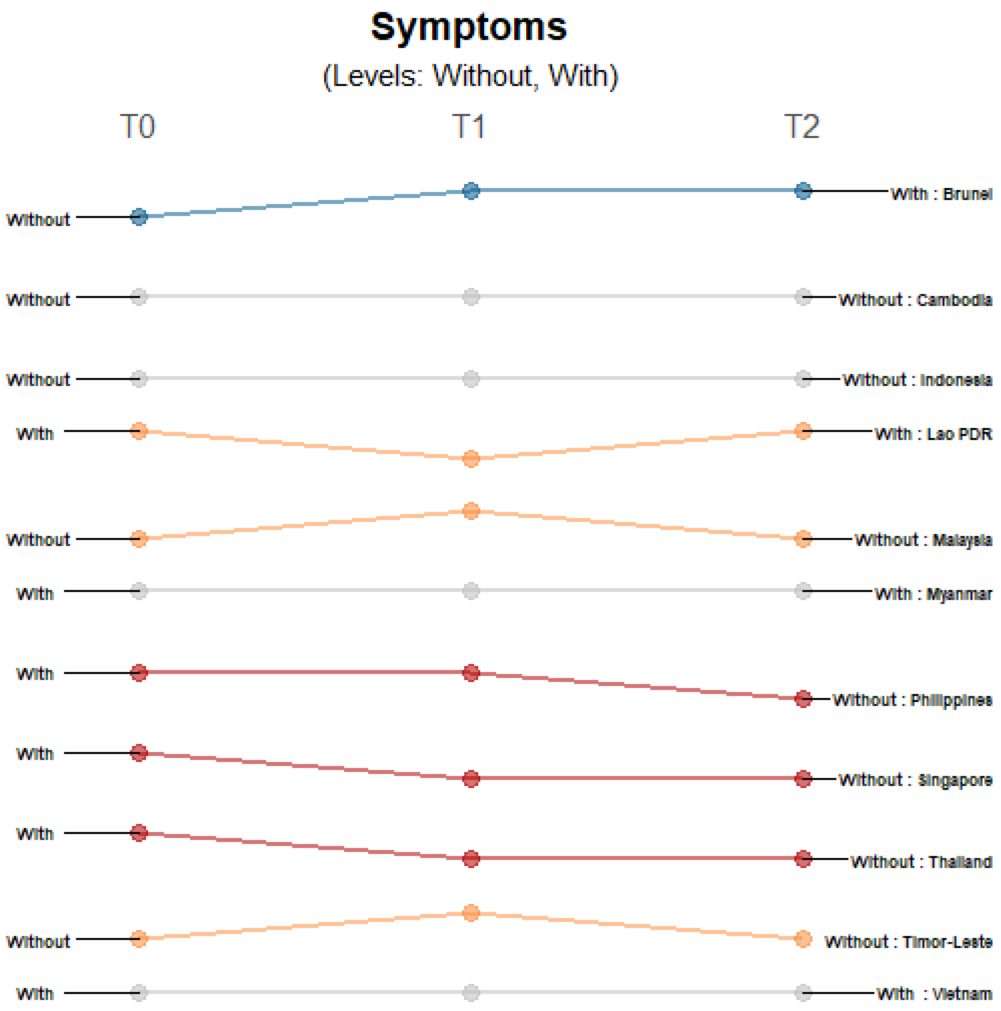


**Supplementary Figure 2. Differences in the level of precision in reporting symptoms**. Only those countries with changes in the level of detail and precision of reporting are highlighted. Each country may shift reporting at any timepoint: at the first reporting of cases' (T0), 'first observed change in reporting' (T1), and 'last observed change in reporting' (T2). Each country may report less precise data indicated by a decreasing slope (red) or more precise data indicated by an increasing slope (blue) consistently over time. Reporting may not be consistent across timepoints with shifts between different levels of precision (yellow) or reporting may not have changed at all during the study period (grey). The levels of precision are indicated for this epidemiological variable. This variable has two levels: without and with.

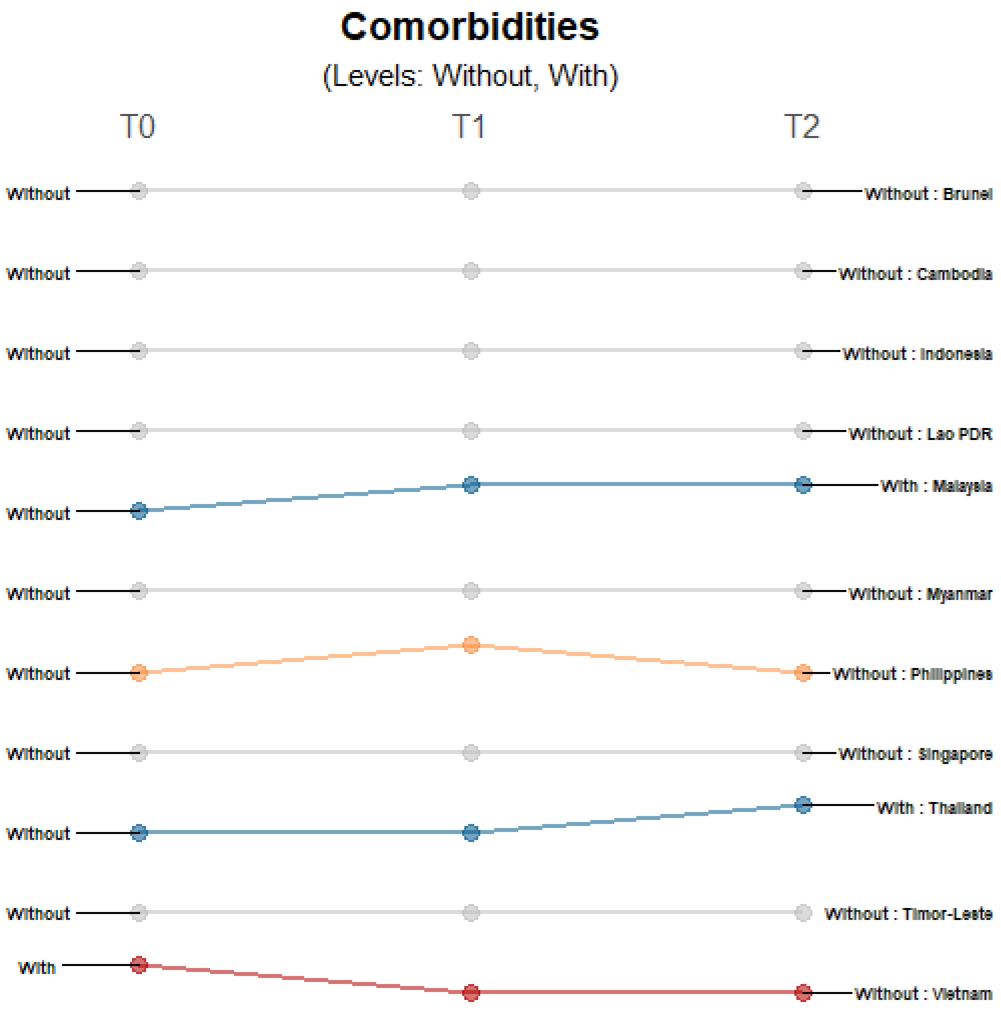


**Supplementary Figure 3. Differences in the level of precision in reporting comorbidities**. Only those countries with changes in the level of detail and precision of reporting are highlighted. Each country may shift reporting at any timepoint: at the first reporting of cases' (T0), 'first observed change in reporting' (T1), and 'last observed change in reporting' (T2). Each country may report less precise data indicated by a decreasing slope (red) or more precise data indicated by an increasing slope (blue) consistently over time. Reporting may not be consistent across timepoints with shifts between different levels of precision (yellow) or reporting may not have changed at all during the study period (grey). The levels of precision are indicated for this epidemiological variable. This variable has two levels: without and with.

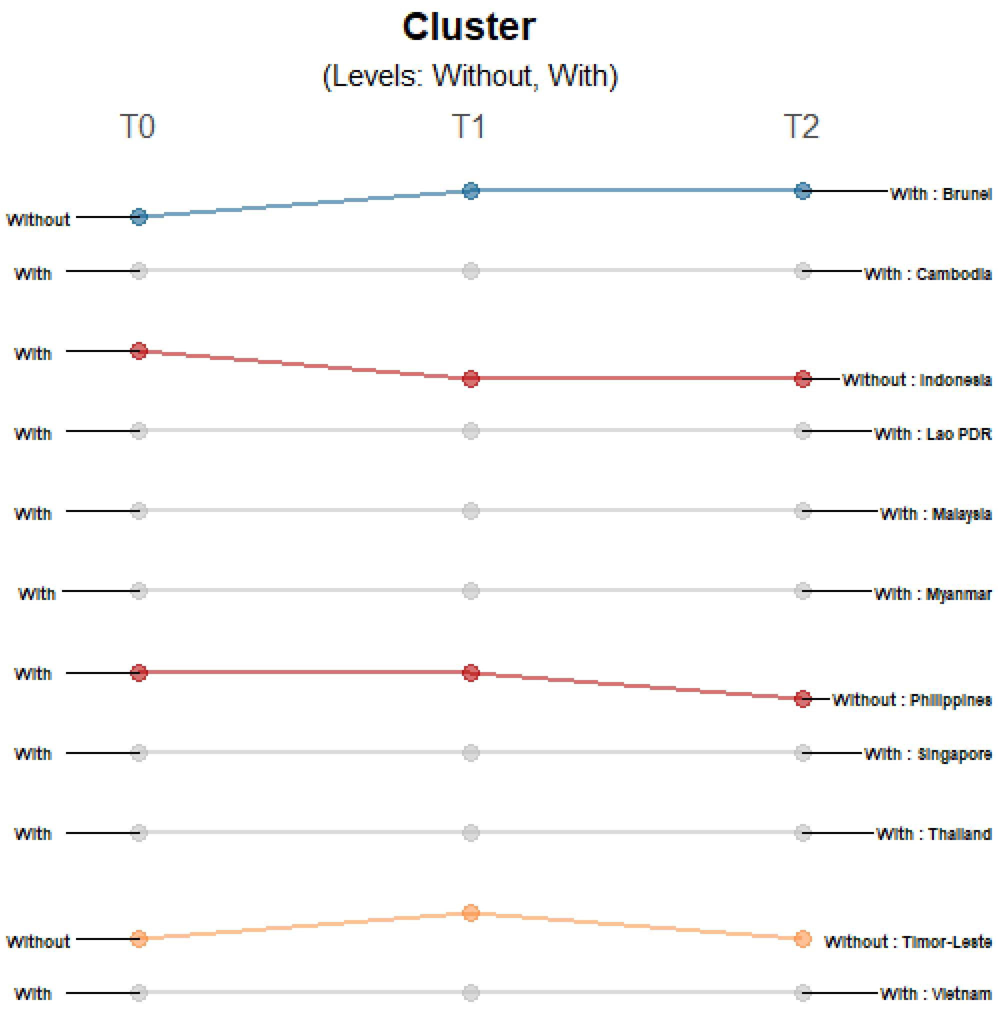


**Supplementary Figure 4. Differences in the level of precision in reporting cluster data**. Only those countries with changes in the level of detail and precision of reporting are highlighted. Each country may shift reporting at any timepoint: at the first reporting of cases' (T0), 'first observed change in reporting' (T1), and 'last observed change in reporting' (T2). Each country may report less precise data indicated by a decreasing slope (red) or more precise data indicated by an increasing slope (blue) consistently over time. Reporting may not be consistent across timepoints with shifts between different levels of precision (yellow) or reporting may not have changed at all during the study period (grey). The levels of precision are indicated for this epidemiological variable. This variable has two levels: without and with.

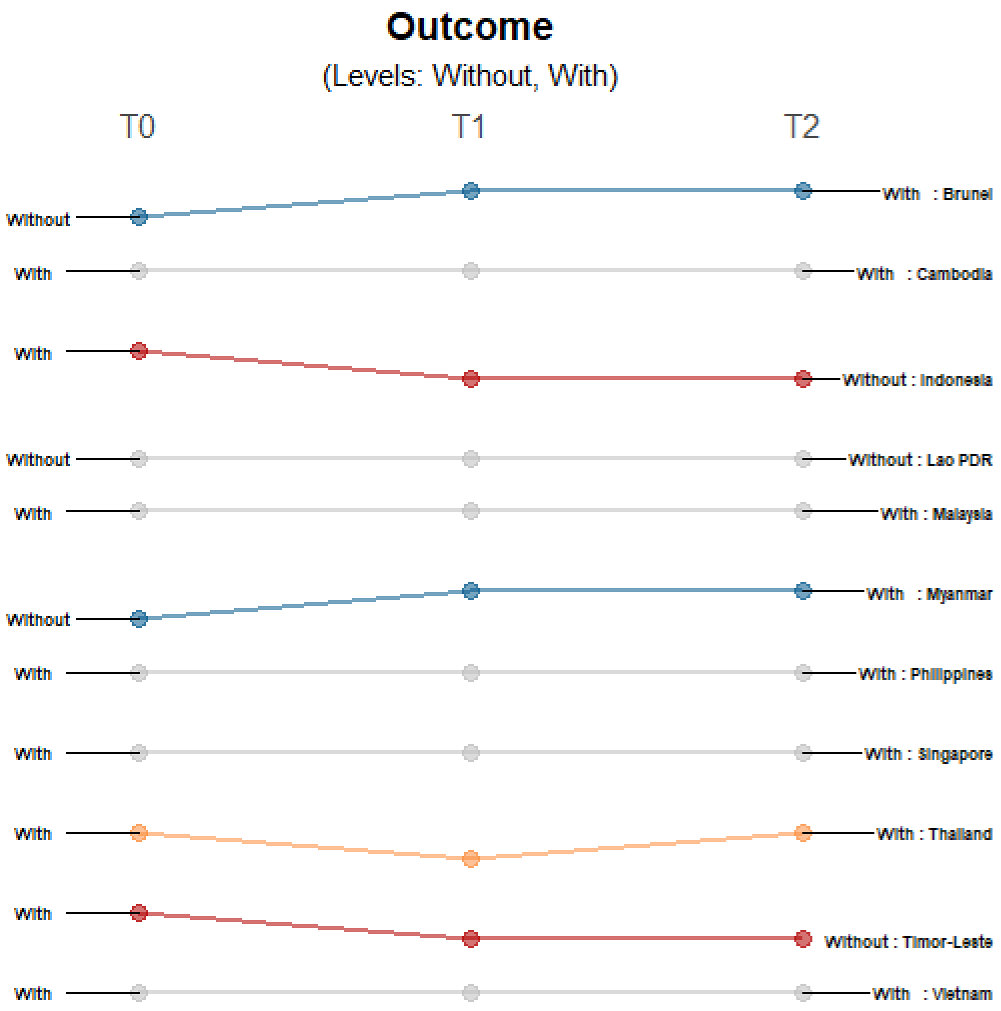


**Supplementary Figure 5. Differences in the level of precision in reporting outcome data**. Only those countries with changes in the level of detail and precision of reporting are highlighted. Each country may shift reporting at any timepoint: at the first reporting of cases' (T0), 'first observed change in reporting' (T1), and 'last observed change in reporting' (T2). Each country may report less precise data indicated by a decreasing slope (red) or more precise data indicated by an increasing slope (blue) consistently over time. Reporting may not be consistent across timepoints with shifts between different levels of precision (yellow) or reporting may not have changed at all during the study period (grey). The levels of precision are indicated for this epidemiological variable. This variable has two levels: without and with.
